# BOLD cerebrovascular reactivity MRI to identify tissue reperfusion failure after EVT in patients with LVO acute stroke

**DOI:** 10.1101/2023.05.12.23289821

**Authors:** Jacopo Bellomo, Martina Sebök, Vittorio Stumpo, Christiaan HB van Niftrik, Darja Meisterhans, Marco Piccirelli, Lars Michels, Beno Reolon, Tilman Schubert, Zsolt Kulcsar, Andreas R Luft, Susanne Wegener, Luca Regli, Jorn Fierstra

**Author notes:** **Corresponding author: Jacopo Bellomo** Department of Neurosurgery, University Hospital Zurich, Zurich, Switzerland Frauenklinikstrasse 10, CH-8091 Zurich. **Authorship contributions:** Data collection and curation: J.B., M.S., V.S., CHB.V., D.M. Analysis of the data and preparing the manuscript draft: J.B., J.F. Revising the manuscript: all authors Supervising the study: J.F., S.W., A.L. All authors approved the final manuscript version.

## Abstract

**Background and purpose:** In acute ischemic stroke due to large-vessel occlusion (LVO), the clinical outcome after endovascular thrombectomy (EVT) is influenced by the extent of autoregulatory hemodynamic impairment and collateral recruitment, which can be derived from blood oxygenation-level dependent cerebrovascular reactivity (BOLD-CVR). BOLD-CVR imaging identifies brain areas influenced by hemodynamic steal. We sought to investigate the presence of steal phenomenon and its relationship to DWI lesions and clinical deficit in the acute phase of ischemic stroke following successful vessel recanalization.

**Methods:** From the prospective longitudinal IMPreST (Interplay of Microcirculation and Plasticity after ischemic Stroke) cohort study, patients with acute ischemic unilateral LVO stroke of the anterior circulation with successful endovascular thrombectomy (EVT; mTICI scale ≥ 2b) and subsequent BOLD-CVR examination were included for this analysis. We analyzed the spatial correlation between brain areas exhibiting BOLD-CVR associated steal phenomenon and DWI infarct lesion as well as the relationship between steal phenomenon and NIHSS score at hospital discharge.

**Results:** Included patients (n=21) exhibited steal phenomenon to different extents, whereas there was only a partial spatial overlap with the DWI lesion (median 18.51%; IQR, 8.44-59.09). The volume of steal phenomenon outside the DWI lesion showed a positive correlation with overall DWI lesion volume and was a significant predictor for the NIHSS score at hospital discharge.

**Conclusions:** Patients with acute ischemic unilateral LVO stroke exhibited hemodynamic steal identified by BOLD-CVR after successful EVT. Steal volume was associated with DWI infarct lesion size and with poor clinical outcome at hospital discharge. BOLD-CVR may further aid in better understanding persisting hemodynamic impairment following reperfusion therapy.

## INTRODUCTION

Acute ischemic large-vessel occlusion (LVO) stroke is characterized by proximal occlusion of a large cerebral vessel (e.g., internal carotid artery or middle cerebral artery) and has a profound hemodynamic impact on distal brain tissue, i.e., hypoperfusion and tissue ischemia.^1^ Hypoperfusion and consecutive tissue damage is a dynamic process in the acute ischemic phase which is influenced by collateral recruitment and autoregulation.^1–4^ Timely restoration of cerebral blood flow by endovascular thrombectomy (EVT) is the most effective maneuver for salvaging ischemic brain tissue that is not already irreversibly lost.^5–8^ However, in some cases no improvement or even worsening of clinical status despite successful vessel recanalization can be observed. It is hypothesized that the mechanisms underlying this reperfusion failure are attributable to remaining macro- and/or microvascular dysfunction and subsequent persisting hemodynamic impairment within the ischemic territory.^9–11^

In recent years, several neuroimaging techniques have been employed investigating microvasculature and hemodynamic tissue responses following acute ischemia and subsequent endovascular revascularization to predict tissue fate.^10,12,13^ Hyper- and hypoperfusion tissue status observed with perfusion-weighted magnetic resonance imaging (MRI) have been both associated with poor clinical outcome. For instance, hypoperfusion despite successful EVT is thought to result from insufficient capillary reflow induced by different mechanisms (e.g. microvascular occlusion with microclots or neutrophils)^13–16^. Hyperperfusion, most likely, comes from persisting altered cerebrovascular autoregulation^13,17^. A better characterization of hemodynamic tissue state in these patients would help to further understand the mechanisms underlying reperfusion failure.

Blood oxygenation-level dependent cerebrovascular reactivity (BOLD-CVR) may be suited as an emerging, clinically applicable, hemodynamic imaging technique capable to evaluate vessel reactivity and remaining vasodilatory reserve as the result of flow redistribution under a controlled hypercapnic challenge.^18–20^ Of particular interest are brain areas exhibiting BOLD-CVR associated steal phenomenon in the post-reperfusion period, indicating persisting severely impaired cerebrovascular autoregulatory loss and insufficient collateral compensatory activation.^21,22^ Recently, we have enabled a clinical infrastructure for advanced MRI investigations in patients presenting with acute ischemic unilateral LVO stroke after reperfusion therapy (IMPreST prospective cohort study, https://www.stroke.uzh.ch/en.html).

We therefore studied the presence of BOLD-CVR identified steal, i.e., a paradoxical BOLD signal drop during hypercapnia^19^, and its association with diffusion-weighted imaging (DWI) lesions, as well as clinical outcome after acute ischemic unilateral LVO stroke following successful endovascular thrombectomy.

## MATERIALS AND METHODS

### Study population

From the prospective IMPreST (Interplay of Microcirculation and Plasticity after ischemic Stroke) longitudinal observational cohort study, we selected all patients with acute ischemic unilateral LVO stroke of the anterior circulation that were successfully treated with EVT (modified Thrombolysis In Cerebral Infarction - mTICI - scale ≥ 2b^23^) and received BOLD-CVR examination. The IMPreST study is a prospective study designed to explore the correlation between different imaging modalities for microcirculation and its association with clinical outcome in patients with acute ischemic unilateral LVO stroke. Inclusion criteria were: (1) ≤ 72 hours first-ever clinical ischemic stroke at hospital admission; (2) occlusion of M1/M2-segment of the middle cerebral artery, and/or intracranial internal carotid artery, and perfusion deficits with cortical involvement; (3) 18 years or above; (4) living independent before stroke (modified Ranking Scale - mRS - ≤ 3^24^); (5) written informed consent of the patient or when the patient is not able to participate in the consenting procedure, the written authorization of an independent doctor who is not involved in the research project to safeguard the interests of the patients (in that case, post-hoc written informed consent of the patient or next of kin had to be obtained). Exclusion criteria were: (1) major cardiac, psychiatric and/or neurological diseases; (2) early seizures; (3) known or suspected non-compliance, drug and/or alcohol abuse; (4) contra-indications for MRI; (5) documented evidence that the patient does not want to participate in any scientific study. Included patients received standard multimodality MRI at predefined time points (i.e., ≤ 72 hours, at day 7±3, at day 90±14) from stroke symptom onset. The exact scanning protocol can be reviewed in the **Supplementary material** section. For this work, we considered only BOLD-CVR and DWI data acquired during the first examination session (i.e., ≤ 72 hours from stroke symptom onset).

### Ethics

The research ethic committee of the Canton Zurich, Switzerland (Kantonale Ethikkommission Zürich; KEK-ZH-NR. 2019-00750) approved the IMPreST prospective observational cohort study. Written informed consent was obtained from each participant before inclusion. The study was conducted in accordance with the ethical standards as laid down in the 1964 Declaration of Helsinki and its later amendments.

### Image acquisition protocol

The imaging study was performed at 3-Tesla Skyra MRI scanner (Siemens Healthineers, Forchheim, Germany) with a 32-channel head matrix coil after the patients have been enrolled in the study. The imaging protocol included diffusion-weighted imaging (DWI) [2D EPI sequence, repetition and echo time (TR/TE) = 2500/75 ms, flip angle = 90°, slice thickness (ST) = 5 mm, *b*-values = B0 and B1000 s/mm^2^], BOLD imaging [2D EPI sequence, TR/TE = 2000/30 ms, flip angle = 85°, bandwidth 2368 Hz/Px, and field of view (FOV) 192×192 mm^2^], 3D T1-MPRAGE imaging [TR/TE = 2200/5.14 ms, flip angle 8°, ST 1 mm, FOV 230×230 mm^2^]. During the BOLD MRI sequence, a standardized carbon dioxide (CO_2_) stimulus was applied using the RespirAct™ (Thornhill Research Institute, Toronto, Canada), that allows for precise CO_2_ end-tidal pressure (P_et_CO_2_) targeting while maintaining normal levels of O_2_ (iso-oxia).^25^ Our standardized CO_2_ protocol consisted of an initial 100 s at the patient-specific resting P_et_CO_2_, after which P_et_CO_2_ was increased of 10 mmHg for 80 s, and a return to resting P_et_CO_2_ for 120 s; P_et_O_2_ was maintained at the patient-specific resting value for the entire duration of the examination.

As specified above, BOLD-CVR imaging and DWI data were collected in a single examination session within 72 hours from stroke symptom onset. The quality of the single BOLD-CVR imaging data was evaluated considering head motion artifacts and the consistency of the CO_2_ stimulus. Specifically, examination were discarded from our analysis if the mean frame-wise displacement between adjacent acquisition volumes was > 2 mm^26^ or if the CO_2_ step change was < 6 mmHg.

### Image processing

Morphological and functional images were first processed singularly to calculate parameter maps. BOLD-CVR maps were obtained according to the previously described Zurich analysis pipeline^27^ using MATLAB2019 (The MathWorks, Inc., Natrick, United States) and SPM12 (Wellcome Trust Centre for Neuroimaging, Institute of Neurology, University College London). BOLD-CVR was calculated voxel-per-voxel as percentage of BOLD signal change divided by the absolute change in P_et_CO_2_ (% ΔBOLD/mmHg). Apparent diffusion coefficient (ADC) maps were automatically calculated from DWI data. All the resulting maps were then co-registered to the individual anatomical T1 space (intra-individual co-registration) using SPM12. Lastly, we extrapolated quantitative values from the co-registered parameter maps for different region of interests (ROIs) - i.e., whole brain (WB), grey matter (GM) and white matter (WM), ipsilateral and contralateral hemisphere, and major vascular territories (anterior cerebral artery, ACA; middle cerebral artery, MCA; posterior cerebral artery, PCA). ROIs of the vascular territories were provided by the recently published atlases by Liu et al.^28^ To better investigate brain areas under steal, we selected for each patient the relevant voxels showing negative response in the BOLD-CVR map. First, we considered all voxels with < 0% BOLD signal change/mmHg CO_2_. Then, using our healthy atlas as reference, a Z-score map^29^ of the BOLD-CVR map was generated and the voxels with Z-score < 2 were excluded. In this way, only those negative voxels that differed significantly from the healthy cohort were considered (**Supplementary Figure 1**). Additionally, for each patient the deep learning-based algorithm by Liu et al. was used to automatically segment stroke lesion from DWI data (DWI infarct stroke lesion).^30^

### Statistical analysis

The statistical analysis of this study was carried out with the statistical program *R studio* (Posit Software, PBC formerly R Studio, version 02.07.2022). First, we described for each patient the mean BOLD-CVR values as well as the volumetric distribution of BOLD-CVR associated steal phenomenon in the different vascular territories and within the DWI infarct stroke lesion. Secondly, we looked at the spatial correlation between BOLD-CVR associated steal phenomenon and DWI infarct lesion by calculating the percentage of tissue exhibiting steal phenomenon that was also included in the infarct lesion. We defined therefore DWI-positive steal phenomenon-positive respectively DWI-negative steal phenomenon-positive brain tissue areas, and we performed a Spearman’s correlation analysis to study the partial correlation between DWI-negative steal phenomenon-positive brain tissue and DWI infarct lesion with symptom-to-MR time (in hour, defined as time between stroke symptom onset and baseline MR study session), age and pre-thrombectomy NIHSS values as confounding variables (covariates). Lastly, we modeled a multivariable linear regression model to study the effect of the presence of DWI-negative steal-positive brain tissue on NIHSS values at discharge with DWI lesion volumes, pre-thrombectomy NIHSS values, and age as confounding variables (covariates).

## RESULTS

Between October 2019 and February 2022, twenty-six patients with acute ischemic unilateral LVO stroke consecutively included in the IMPreST prospective study received BOLD-CVR examination within 72 hours from stroke symptom onset. Of these, 5 patients were excluded due to excessive head movement during MR examination. Twenty-one patients were included in analysis (**Figure 1**) and their baseline characteristics can be reviewed in **Table 1**.

**Figure 1.**
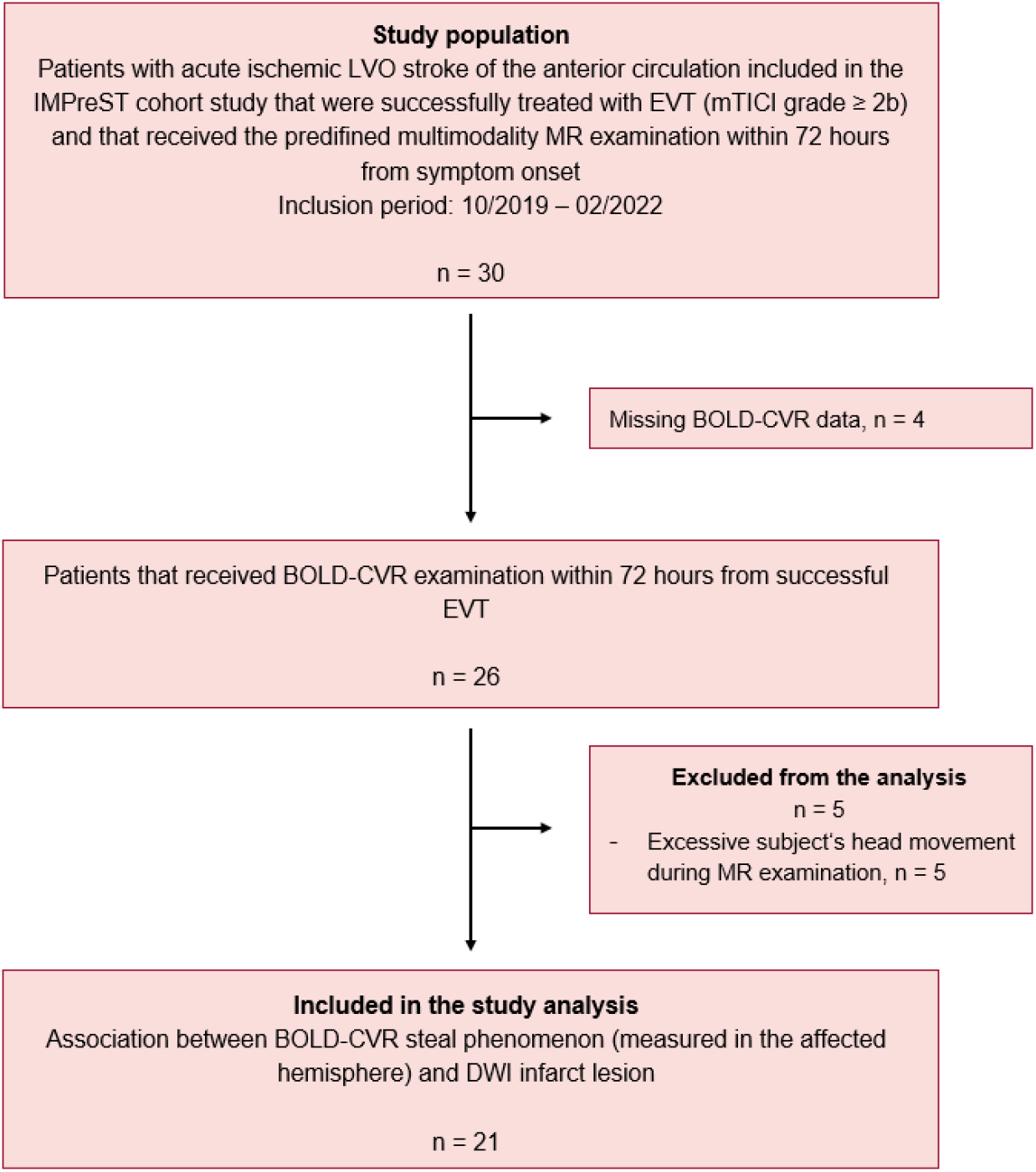
Study flow chart. LVO indicates large-vessel occlusion stroke; IMPreST, Interplay of Microcirculation and Plasticity after ischemic Stroke; EVT, endovascular thrombectomy; mTICI, modified Thrombolysis in Cerebral Infarction; MR, magnetic resonance; BOLD-CVR, blood oxygen level dependent cerebrovascular reactivity; DWI, diffusion-weighted imaging.

**Table 1.** Baseline characteristics. ICA indicates internal carotid artery; MCA, middle carotid artery; NIHSS, National Institutes of Health Stroke Scale; mRS, modified Ranking Scale; mTICI, modified Thrombolysis in Cerebral Infarction; IVT, intravenous thrombolysis; EVT, endovascular thrombectomy; MR, magnetic resonance; DWI, diffusion-weighted imaging.

| Baseline demographics | All (n = 21) |
| --- | --- |
| **Age** |  |
| Mean ± SD | 69 ± 15 |
| **Year group** |  |
| <50 | 3 (14%) |
| 50-70 | 8 (38%) |
| >70 | 10 (48%) |
| **Sex** |  |
| M | 9 (43%) |
| F | 12 (57%) |
| **Diseased vessel** |  |
| ICA | 2 (10%) |
| MCA (M1/M2 segment) | 11 (52%) |
| Both | 8 (38%) |
| **Clinical score at hospital admission** |  |
| **NIHSS** |  |
| Median [IQR] | 10.00 [8.00-16.00] |
| **mRS** |  |
| Median [IQR] | 4.00 [3.00-5.00] |
| **Comorbidities** |  |
| Atrial fibrillation | 4 (20%) |
| Smoking history | 5 (24%) |
| Hypertension | 13 (55%) |
| Dyslipidemia | 8 (40%) |
| Obesity | 3 (15%) |
| Diabetes | 1 (5%) |
| **Acute reperfusion therapy** |  |
| IVT | 10 (50%) |
| EVT | 21 (100%) |
| **Symptom-to-needle time** |  |
| <4.5 hours | 12 (57%) |
| >4.5 hours | 2 (10%) |
| Wake-up stroke | 7 (33%) |
| **mTICI grade** |  |
| 2b | 5 (24%) |
| 2c | 3 (14%) |
| 3 | 13 (62%) |
| **Symptom-to-MR time** |  |
| <24 hours | 3 (30%) |
| 24-48 hours | 11 (40%) |
| 48-72 hours | 7 (30%) |
| **DWI infarct lesion [mL]** |  |
| Median [IQR] | 26.30 [14.58-52.84] |

In the MR examination session after successful reperfusion therapy, we found high variability of the presence of steal phenomenon in the affected MCA and ACA territory of the included patients, with a median volume of 21.58 mL (IQR, 7.68-64.11). **Table 2** summarizes the volumetric distribution of BOLD-CVR steal for all the included patients. Interestingly, steal-affected regions were heterogeneously distributed both within and outside the DWI lesion. On average, only 18.51% (INR, 8.44-59.09) of DWI lesion overlapped the steal area; respectively only 26.31% (IQR, 13.84-39.47) of steal volume was part of the DWI lesion (**Supplementary Figure 2**). Within the DWI lesion, on average, severely impaired BOLD-CVR values were observed (mean ± SD; 0.042 ± 0.058 ΔBOLD%/mmHg). A Spearman’s correlation analysis showed a positive partial correlation (correlation coefficient 0.64; SE 0.20; *P* value 0.004) between DWI lesion volume and BOLD-CVR steal volume found outside the stroke lesion after correcting for possible confounding variables (age, symptom-to-MR time, and NIHSS baseline values).

**Table 2.** Distribution of BOLD-CVR associated steal phenomenon in the affected hemisphere. BOLD-CVR values are reported with unit [% ΔBOLD/mmHg]; volumes are reported with unit [mL]; BOLD-CVR indicates blood oxygen level dependent cerebrovascular reactivity; SP, steal phenomenon; GM, grey matter; WM, white matter; ACA, anterior cerebral artery; MCA, middle cerebral artery; PCA, posterior cerebral artery.

| Subject | Mean BOLD-CVR in MCA territory | Mean BOLD-CVR in ACA territory | Mean ± SD BOLD-CVR in DWI lesion | DWI infarct lesion volume | SP volume in ACA + MCA territory | SP volume in DWI lesion | SP volume outside DWI lesion |
| --- | --- | --- | --- | --- | --- | --- | --- |
| 1 | 0.07 | 0.11 | 0.04 ± 0.01 | 10.18 | 14.94 | 3.84 | 11.09 |
| 2 | 0.09 | 0.07 | 0.02 ± 0.01 | 14.58 | 42.64 | 6.42 | 36.22 |
| 3 | 0.08 | 0.14 | 0.06 ± 0.01 | 47.15 | 4.40 | 0.70 | 3.70 |
| 4 | 0.11 | 0.09 | 0.08 ± 0.01 | 2.02 | 2.71 | 0.06 | 2.65 |
| 5 | 0.06 | 0.08 | 0.01 ± 0.01 | 97.12 | 189.76 | 73.84 | 115.92 |
| 6 | -0.03 | -0.01 | -0.03 ± 0.02 | 117.29 | 174.03 | 73.75 | 100.28 |
| 7 | 0.08 | 0.06 | -0.03 ± 0.02 | 72.13 | 129.37 | 45.88 | 83.49 |
| 8 | 0.08 | 0.06 | 0.06 ± 0.01 | 26.30 | 8.77 | 1.24 | 7.54 |
| 9 | 0.12 | 0.08 | 0.04 ± 0.01 | 13.89 | 4.07 | 1.34 | 2.73 |
| 10 | 0.14 | 0.12 | 0.08 ± 0.01 | 4.29 | 2.59 | 0.00 | 2.60 |
| 11 | 0.19 | 0.23 | 0.09 ± 0.02 | 18.00 | 21.58 | 2.79 | 18.78 |
| 12 | 0.15 | 0.20 | 0.03 ± 0.03 | 36.41 | 42.69 | 19.34 | 23.35 |
| 13 | 0.08 | 0.08 | -0.01 ± 0.01 | 89.57 | 83.99 | 51.79 | 32.20 |
| 14 | 0.06 | 0.00 | 0.03 ± 0.01 | 17.74 | 25.79 | 3.28 | 22.51 |
| 15 | 0.20 | 0.14 | 0.17 ± 0.02 | 1.33 | 0.36 | 0.00 | 0.36 |
| 16 | 0.06 | 0.15 | -0.05 ± 0.01 | 52.84 | 113.19 | 46.55 | 66.64 |
| 17 | 0.04 | 0.05 | -0.04 ± 0.02 | 23.30 | 57.50 | 17.92 | 39.57 |
| 18 | 0.13 | 0.10 | 0.08 ± 0.01 | 30.78 | 18.49 | 4.97 | 13.52 |
| 19 | 0.18 | 0.13 | 0.13 ± 0.02 | 16.18 | 9.51 | 2.11 | 7.40 |
| 20 | 0.11 | 0.10 | 0.05 ± 0.01 | 80.20 | 33.94 | 21.05 | 12.89 |
| 21 | 0.14 | 0.12 | 0.08 ± 0.01 | 30.30 | 4.08 | 0.59 | 3.49 |

At hospital discharge the median NIHSS score value was 2.00 (IQR, 1.00-7.25). Five-teen patients had a NIHSS score equal or less than 3 (median, min-max; 1, 0-3), the remaining six patients more than 3 (median, min-max; 9.50, 7.00-18.00). A comparison of NIHSS values at hospital admission (pre-thrombectomy) and at hospital discharge are presented in **Figure 2**.

**Figure 2.**
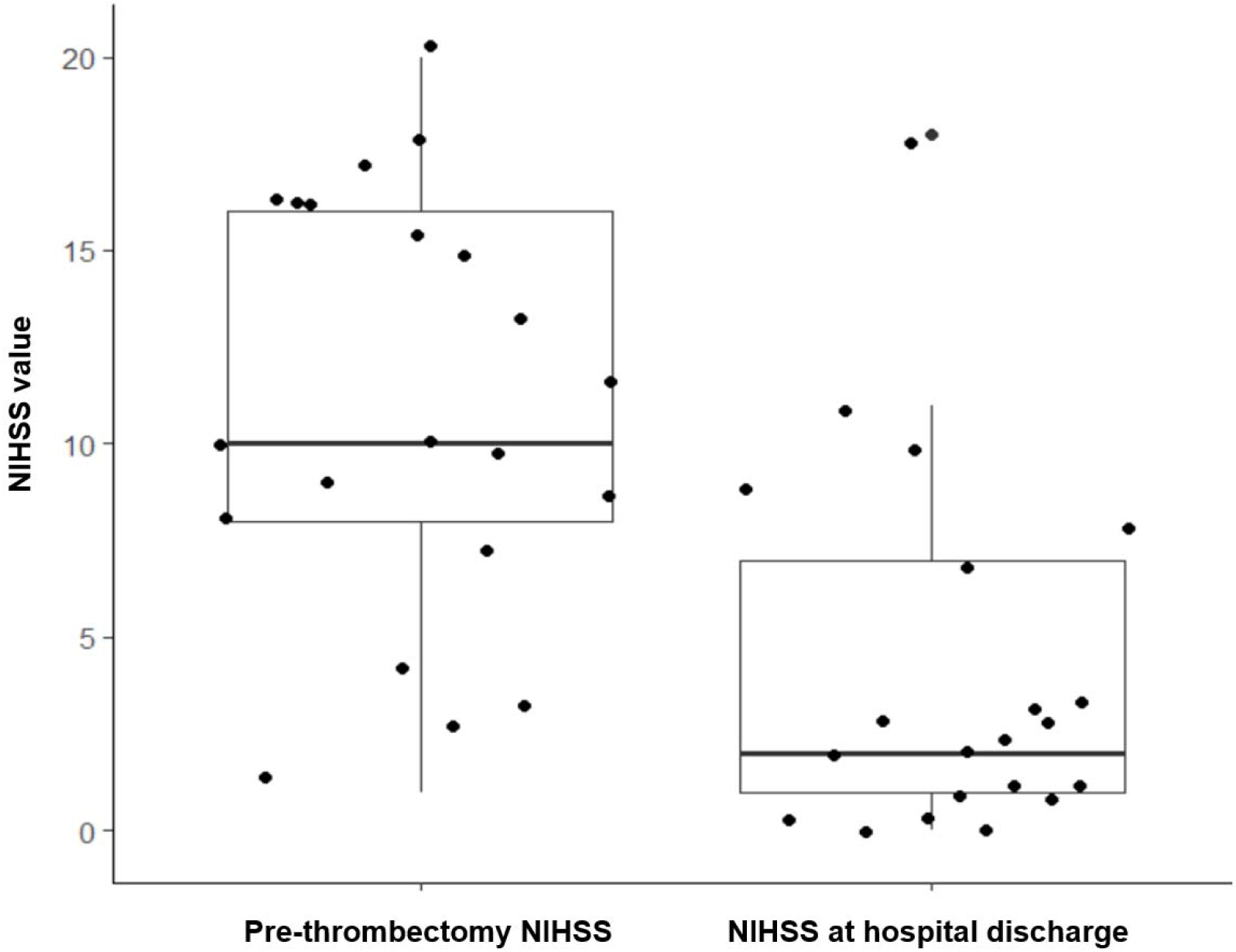
Boxplot of NIHSS score measurements over time. NIHSS indicates National Institutes of Health Stroke Scale.

A multivariable linear regression model (**Table 3**) showed that the volume of BOLD-CVR defined steal found outside the DWI lesion was a significant predictor for the NIHSS score at discharge (regression coefficient β 0.07, SE 0.03, *P* value 0.034) after correcting for DWI lesion size, pre-thrombectomy NIHSS values, and age (adjusted R^2^ 0.64, *P* value <0.001). In **Figure 3**, BOLD-CVR and DWI findings of four example patients are presented.

**Table 3.** Results of the linear regression model of the association between BOLD-CVR associated steal phenomenon volume outside the infarct lesion and NIHSS score value at hospital discharge. β indicates the regression coefficient; BOLD-CVR, blood oxygen level dependent cerebrovascular reactivity; SP, steal phenomenon; DWI, diffusion-weighted imaging; NIHSS, National Institutes of Health Stroke Scale.

| Variable | $\beta$ | 95% confidence interval (95-CI) | P value |
| --- | --- | --- | --- |
| Intercept | 2.34 | -5.57 to 10.25 | 0.540 |
| Age | -0.06 | -0.17 to 0.04 | 0.223 |
| BOLD-CVR associated SP outside the infarct lesion (mL) | 0.07 | 0.01 to 0.14 | **0.034\*** |
| DWI infarct lesion volume (mL) | 0.03 | -0.03 to 0.10 | 0.260 |
| NIHSS at hospital admission | 0.23 | -0.06 to 0.53 | 0.116 |

**Figure 3.**
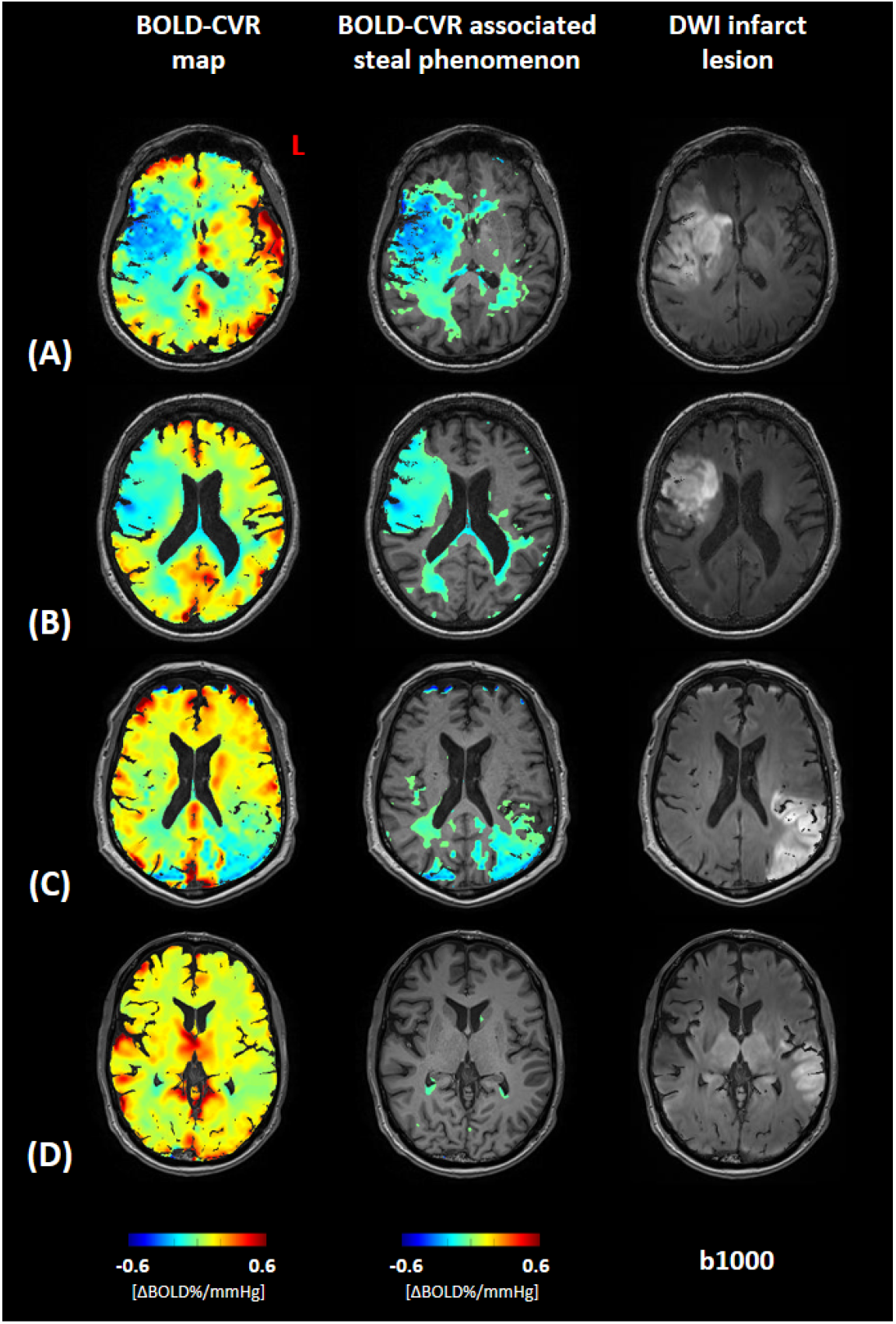
Association between BOLD-CVR associated steal phenomenon and DWI infarct lesion. Blood oxygen level dependent cerebrovascular reactivity (BOLD-CVR) and trace DWI (diffusion-weighted imaging) map of four illustrative examples are presented. For each patient a whole-brain BOLD-CVR map, a BOLD-CVR showing only steal phenomenon tissue areas, and trace DWI map are depicted. **A**, male subject (age range: 66-70 years) showing 97.12 mL infarct lesion in the middle cerebral artery (MCA) territory right, showing a total of 189.76 mL steal phenomenon, 73.84 mL within and 115.52 mL outside the infarct lesion. NIHSS 16 at hospital admission, NIHSS 9 at hospital discharge. **B**, female subject (age range: 66-70 years) showing 52.84 mL infarct lesion in the MCA territory right, showing a total of 113.19 mL steal phenomenon, 46.55 mL within and 66.64 mL outside the infarct lesion. NIHSS 12 at hospital admission, NIHSS 10 at hospital discharge. **C**, male subject (age range: 71-75 years) showing 89.57 mL infarct lesion in the MCA territory left, showing a total of 83.99 mL steal phenomenon, 51.79 mL within and 32.20 mL outside the infarct lesion. NIHSS 10 at hospital admission, NIHSS 1 at hospital discharge. **D**, male subject (age range: 66-70 years) showing 47.15 mL infarct lesion in the MCA territory left, showing a total of 4.40 mL steal phenomenon, 0.70 mL within and 3.70 mL outside the infarct lesion. NIHSS 3 at hospital admission, NIHSS 1 at hospital discharge.

## DISCUSSION

Our study shows that BOLD-CVR associated steal phenomenon can be detected in the affected hemisphere of patients with acute ischemic unilateral LVO stroke despite successful EVT, which may indicate reperfusion failure. Brain areas with steal phenomenon only showed a partial spatial agreement with DWI infarct lesions, whereas steal phenomenon was also observed in brain tissue outside the DWI lesion. The brain tissue volume exhibiting steal phenomenon outside the infarct – DWI – lesion correlated strongly with the volume of DWI derived infarct lesion and was a significant predictor for a poorer clinical status at hospital discharge.

To the best of our knowledge, we could not identify similar studies measuring CVR after acute reperfusion therapy in a similar cohort of patients, and therefore a direct comparison of our results with already existing findings is not possible to date. However, some clinical studies reported the findings observed with other imaging techniques.^31–35^ A dynamic growth of DWI lesion after revascularization procedures has been observed reflecting reperfusion failure.^31,35^ Infarct growth contributes to the final infarct volume, correlates with clinical outcome and has been associated with DWI lesion size pre-EVT as well as mTICI grade < 2b and hypoperfusion post-EVT.^31^ These findings enhance the importance of identifying prognostic factors that are associated with reperfusion failure and can therefore predict infarct lesion evolution after EVT. A recent work of Potreck et al.^33^ studied tissue response after successful EVT with combined perfusion and permeability MR imaging and showed distinct post-reperfusion pathophysiological tissue responses that were associated with different clinical courses. In general, hypoperfusion occurred more often in patients with unfavourable clinical outcome compared to hyperperfusion or unchanged perfusion. BOLD-CVR imaging, thanks to its capability to study vessel reactivity, offers the potential to provide additional information about microvascular functionality. We focused our analysis on brain regions exhibiting BOLD-CVR identified steal because we thought this could represent irreversible tissue damage and/or brain areas subjected to reperfusion failure. Based on the current knowledge on the pathophysiological mechanisms of reperfusion failure^9,14,16^, we hypothesized that steal phenomenon could reflect on of the following: (1) exhausted regional vasodilatory reserve induced by microvascular occlusion with microclots, microvascular occlusion with recruited neutrophils, and/or vessel constriction; (2) loss of cerebrovascular autoregulation. Most of the knowledge about reperfusion failure mechanisms came from pre-clinical studies. Recently, Binder et al.^36^ investigated the role of leptomeningeal collaterals in reperfusion failure in a rodent stroke model and reported a loss of vascular tone (i.e., loss of autoregulation) in the distal MCA branch segments after reperfusion which was associated with a worse clinical outcome.

In our study, we observed a partial spatial agreement between DWI infarct lesion and BOLD-CVR identified steal. We can distinguish three different tissue areas: DWI-positive steal-negative brain tissue, DWI-positive steal-positive brain tissue, and DWI-negative steal-positive brain tissue. In the DWI infarct lesion (“DWI-positive”), both steal phenomenon and impaired, but maintained BOLD-CVR values were observed. Two possible explanations to this finding seem plausible: (1) infarcted necrotic tissue included in the DWI lesion represents tissue with disrupted vessels (i.e., no cerebrovascular reactivity) and therefore shows BOLD-CVR around zero with either slightly positive or slightly negative values^37^; (2) a part of the DWI lesion represents cerebral tissue not yet irreversible damaged that shows maintained cerebrovascular reactivity after successful reperfusion^35,38^. Of major clinical relevance we observed tissue exhibiting a steal phenomenon that was not included in the DWI lesion (“DWI-negative”), which could indicate tissue not irreversibly damaged but with severely impaired hemodynamic characteristics. We found a positive correlation of this DWI-negative steal-positive brain tissue with DWI infarct lesion size and a significant association with higher NIHSS clinical score at hospital discharge. These findings indirectly support what we assumed in the beginning, namely, that the steal phenomenon does not reflect just infarcted tissue and that the identified DWI-negative steal-positive region could depict brain tissue with microvascular dysfunction (i.e., reperfusion failure) that could evolve to an irreversibly infarcted area.

### Future directions

Our preliminary findings highlight the potential of BOLD-CVR imaging as a novel technique to characterize brain tissue hemodynamic responses after EVT. Future research, however, is needed to further understand the link between the observed BOLD-CVR steal phenomenon and reperfusion failure and its association with clinical outcome. A correlation analysis between BOLD-CVR findings and MR perfusion parameters (i.e., cerebral blood flow, CBF, cerebral blood volume, CBV, mean transit time, MTT, time-to-maximum, T_max_) could be performed to confirm our hypothesis of the mechanisms behind steal phenomenon observed in reperfused tissue (i.e., both loss of autoregulation with associated hyperperfusion and microvascular occlusion/constriction with associated hypoperfusion). In addition, a retrospective analysis to identify which pre-thrombectomy clinical/imaging factors are associated with the occurrence of steal phenomenon could be considered.

### Strength and limitations

This work represents a preliminary analysis of BOLD-CVR findings in a well-defined cohort of patients with acute ischemic unilateral LVO stroke. The investigated imaging modalities (BOLD-CVR and DWI) were acquired within the same examination session and therefore high intra-subject comparability between imaging data can be assumed. However, this study has some limitations. First, the small sample size of the studied cohort limited the generalizability of our analysis and therefore further studies including more patients are necessary to confirm our observations. Second, to avoid unethical diagnostic delay, our study did not provide imaging data about tissue state before reperfusion therapy. Therefore, the clinical meaning of BOLD-CVR identified steal before EVT is unknown. Third, we created a steal phenomenon mask from the BOLD-CVR magnitude maps selecting all the voxel exhibiting a negative response to the hypercapnic stimulus. However, the clinical relevance of this steal phenomenon mask has not yet been investigated and other definitions based on different thresholds respectively considering also more technical imaging aspects (e.g., contrast-to-noise ratio) should be also evaluated in the future.

## CONCLUSIONS

Patients with unilateral acute ischemic large-vessel occlusion stroke exhibit BOLD-CVR associated steal in the early phase despite successful EVT. Steal volume was associated with DWI lesion size, and with poor clinical outcome at hospital discharge. The BOLD-CVR identified steal phenomenon may provide a better understanding of persisting hemodynamic impairment following reperfusion therapy.

## Supporting information

The exact scanning protocol can be reviewed in the Supplementary material section.

## Data Availability

All data are available under request by the corrisponding author.

## Abbreviations

LVO: large-vessel occlusion
BOLD: blood oxygen-level dependent
MRI: magnetic resonance imaging
CVR: cerebrovascular reactivity
DWI: diffusion-weighted imaging
EVT: endovascular thrombectomy
NIHSS: National Institutes of Health Stroke Scale
mTICI: modified Thrombolysis In Cerebral Infarction
IMPreST: Interplay of Microcirculation and Plasticity after ischemic Stroke
SD: standard deviation
SE: standard error

## Supplemental Material

Figures S1-S2

Supplemental methods

