## Supplementary material for "BOLD cerebrovascular reactivity MRI to identify tissue reperfusion failure after EVT in patients with LVO acute stroke": The exact scanning protocol can be reviewed in the Supplementary material section.

### SUPPLEMENTAL MATERIAL

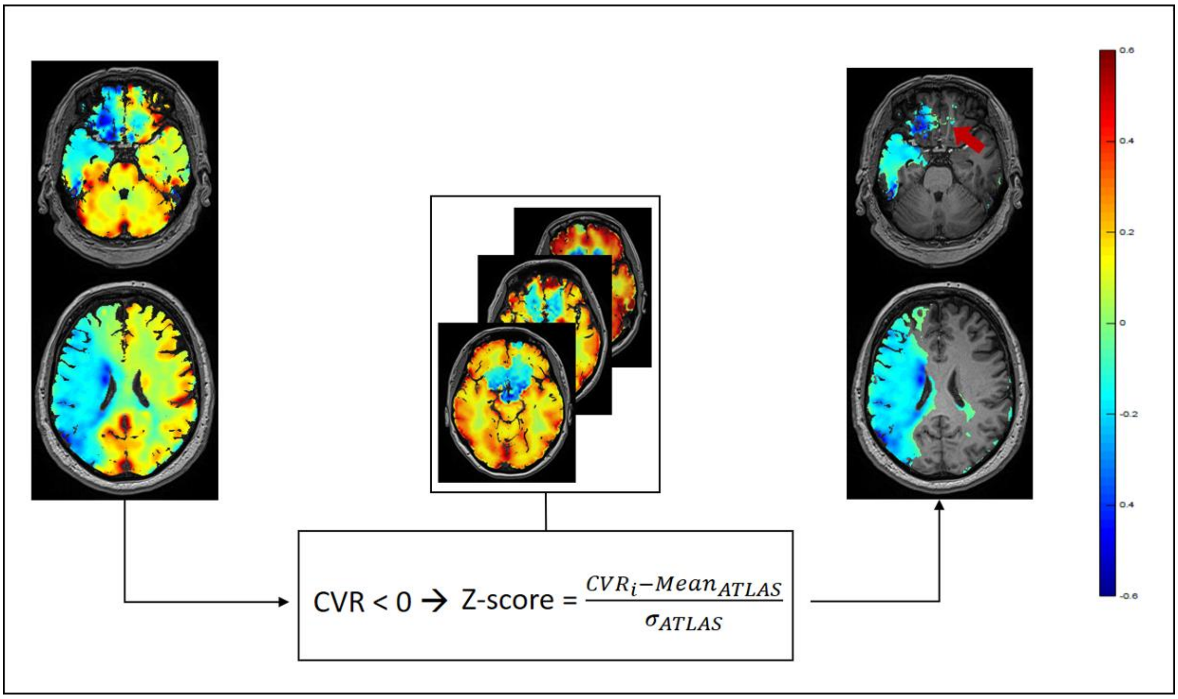


**Supplementary Figure 1. BOLD-CVR associated steal phenomenon mask generation.**

For each patient the relevant voxels showing negative response in the blood oxygen level dependent cerebrovascular reactivity (BOLD-CVR) map were selected. First, we considered all voxels with < 0% BOLD signal change/mmHg $CO_2$. Then, using our healthy atlas as reference, a Z-score map of the CVR map was generated and the voxels with Z-score < 2 were excluded. In this way, only those negative voxels that differed significantly from the healthy cohort (e.g., voxels with artefact-related negative BOLD signal in the frontobasal brain region, red arrow) were considered.

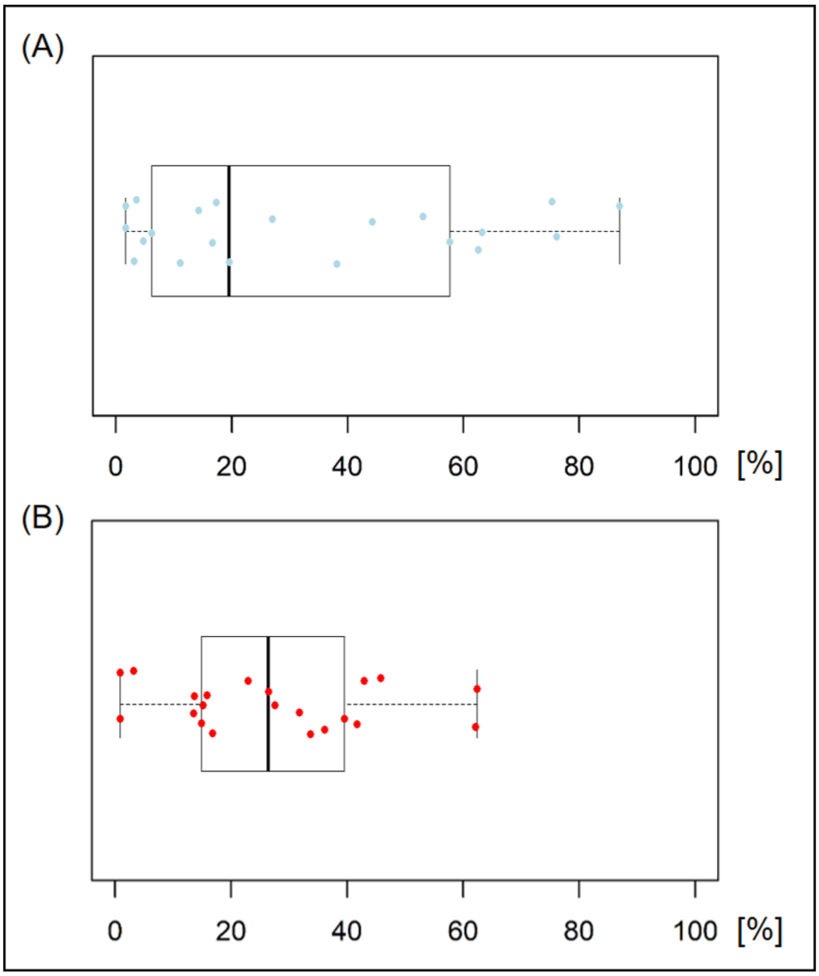


**Supplementary Figure 2. Spatial correlation between BOLD-CVR associated steal phenomenon and DWI infarct lesion.**

BOLD-CVR indicates blood oxygen level dependent cerebrovascular reactivity. **A**, a boxplot of the percentage of DWI infarct lesion included in the BOLD-CVR associated steal phenomenon; **B**, a boxplot of the percentage of BOLD-CVR associated steal phenomenon included in the DWI infarct lesion.

| \\NRA\Schädel_32ch\Ischämie\CRPPstroke20191228_32ch_New_Patients6-\CO2_bold * |
|---|
| TA: 6:46 min Coil Selection: Auto Voxel Size: 3.0×3.0×3.0 mm³ Acc:: 2 Rel. SNR: 1.00 |

### Properties

| | |
|---|---|
| Start measurement without further preparation | On |
| Wait for User to Start | Off |
| Start measurements | Single Measurement |
| Prio Recon | Off |
| Auto Open Inline Display | Off |
| Auto Close Inline Display | Off |
| Load Images to MR View&GO | On |
| Auto Store Images | On |
| Load Images to Stamp Segments | Off |
| Load Images to Graphic Segments | Off |
| Graphic segment | Default |
| Inline Movie | Off |

### Routine

| | |
|---|---|
| Slice Group | 1 |
| Slices | 35 |
| Distance Factor | 10 % |
| Position | R0.8 P5.9 H10.6 mm |
| Orientation | T > C-16.5 > S0.6 |
| Phase Encoding Dir. | A >> P |
| Phase Oversampling | 0 % |
| FoV Read | 192 mm |
| FoV Phase | 100.0 % |
| Slice Thickness | 3.0 mm |
| TR | 2000.0 ms |
| TE | 30.00 ms |
| Averages | 1 |
| Concatenations | 1 |
| AutoAlign | Head > Brain |
| Coil Elements | HEA;HEP |

### Contrast - Common

| | |
|---|---|
| TR | 2000.0 ms |
| TE | 30.00 ms |
| MTC | Off |
| Flip Angle | 85 deg |
| Fat-Water Contrast | Fat Saturation |
| Reconstruction | Magnitude |

### Contrast - Dynamic

| | |
|---|---|
| Dynamic Mode | Standard |
| Measurements | 200 |
| Delay in TR | 0.00 ms |

### Resolution - Common

| | |
|---|---|
| FoV Read | 192 mm |
| FoV Phase | 100.0 % |
| Slice Thickness | 3.0 mm |
| Base Resolution | 64 |
| Phase Resolution | 100 % |
| Interpolation | Off |

### Resolution - Acceleration

| | |
|---|---|
| Acceleration mode | GRAPPA |
| Reference Scans | EPI/Separate |
| Acceleration Factor PE | 2 |
| Reference Lines PE | 32 |
| Phase Partial Fourier | Off |

### Resolution - Filter

| | |
|---|---|
| Raw Filter | Off |
| Elliptical Filter | On |
| Hamming | Off |
| Distortion Correction | Off |
| Normalize | Prescan |

### Geometry - Common

| | |
|---|---|
| Slice Group | 1 |
| Slices | 35 |
| Distance Factor | 10 % |
| Position | R0.8 P5.9 H10.6 mm |
| Orientation | T > C-16.5 > S0.6 |
| Phase Encoding Dir. | A >> P |
| Phase Oversampling | 0 % |
| FoV Read | 192 mm |
| FoV Phase | 100.0 % |
| Slice Thickness | 3.0 mm |
| TR | 2000.0 ms |
| Multi-Slice Mode | Interleaved |
| Series | Interleaved |
| Concatenations | 1 |

### Geometry - AutoAlign

| | |
|---|---|
| Slice Group | 1 |
| Position | R0.8 P5.9 H10.6 mm |
| Orientation | T > C-16.5 > S0.6 |
| Phase Encoding Dir. | A >> P |
| AutoAlign | Head > Brain |
| Initial Position | R0.8 P5.9 H10.6 |
| R | 0.8 mm |
| P | 5.9 mm |
| H | 10.6 mm |
| Initial Orientation | T > C |
| T > C | -16.50 |
| > S | 0.60 |
| Initial Rotation | -1.12 deg |

### Geometry - Saturation

| | |
|---|---|
| Special Saturation | None |

### Geometry - Tim Planning Suite

| | |
|---|---|
| Set-n-Go Protocol | Off |
| Table Position | 0 mm |
| Table Position | H |
| Inline Composing | Off |

### System - Miscellaneous

| | |
|---|---|
| Coil Selection | Auto Coil Select |
| MSMA | S - C - T |
| Sagittal | R >> L |
| Coronal | A >> P |
| Transversal | F >> H |
| Coil Combination | Adaptive Combine |
| Matrix Optimization | Off |

### System - Adjustments

| | |
|---|---|
| Adjustment Strategy | Standard |
| B0 Shim | Standard |
| B1 Shim | TrueForm |
| Adjustment Tolerance | Auto |

### System - Adjustments

| | |
|---|---|
| Adjust with Body Coil | Off |
| Confirm Frequency | Never |
| Assume Silicone | Off |

### System - Adjust Volume

| | |
|---|---|
| Position | R0.8 P5.9 H10.6 mm |
| Orientation | T > C-16.5 > S0.6 |
| Rotation | -1.12 deg |
| A >> P | 192 mm |
| R >> L | 192 mm |
| F >> H | 116 mm |
| Reset | Off |

### System - Tx/Rx

| | |
|---|---|
| Frequency 1H | 123.256229 MHz |
| ? Ref. Amplitude 1H | 0.000 V |
| Reset | Off |
| Correction Factor | 1.00 |
| Image Scaling | 1.000 |

### Physio - Signal

| | |
|---|---|
| 1st Signal/Mode | None |
| TR | 2000.0 ms |
| Concatenations | 1 |

### BOLD

| | |
|---|---|
| GLM Statistics | Off |
| Ignore Meas. at Start | 0 |
| Ignore After Transition | 0 |
| Model Transition States | On |
| Temp. Highpass Filter | On |
| Threshold | 2.50 |
| Paradigm Size | 66 |
| Meas[1] | Active |
| Meas[2] | Active |
| Meas[3] | Active |
| Meas[4] | Active |
| Meas[5] | Active |
| Meas[6] | Active |
| Meas[7] | Active |
| Meas[8] | Active |
| Meas[9] | Active |
| Meas[10] | Active |
| Meas[11] | Active |
| Meas[12] | Active |
| Meas[13] | Active |
| Meas[14] | Active |
| Meas[15] | Active |
| Meas[16] | Active |
| Meas[17] | Active |
| Meas[18] | Active |
| Meas[19] | Active |
| Meas[20] | Active |
| Meas[21] | Active |
| Meas[22] | Active |
| Meas[23] | Ignore |
| Meas[24] | Ignore |
| Meas[25] | Ignore |
| Meas[26] | Ignore |
| Meas[27] | Ignore |
| Meas[28] | Ignore |
| Meas[29] | Ignore |
| Meas[30] | Ignore |
| Meas[31] | Ignore |

### BOLD

| | |
|---|---|
| Meas[32] | Ignore |
| Meas[33] | Ignore |
| Meas[34] | Ignore |
| Meas[35] | Ignore |
| Meas[36] | Ignore |
| Meas[37] | Ignore |
| Meas[38] | Ignore |
| Meas[39] | Ignore |
| Meas[40] | Ignore |
| Meas[41] | Ignore |
| Meas[42] | Ignore |
| Meas[43] | Ignore |
| Meas[44] | Ignore |
| Meas[45] | Active |
| Meas[46] | Active |
| Meas[47] | Active |
| Meas[48] | Active |
| Meas[49] | Active |
| Meas[50] | Active |
| Meas[51] | Active |
| Meas[52] | Active |
| Meas[53] | Active |
| Meas[54] | Active |
| Meas[55] | Active |
| Meas[56] | Active |
| Meas[57] | Active |
| Meas[58] | Active |
| Meas[59] | Active |
| Meas[60] | Active |
| Meas[61] | Active |
| Meas[62] | Active |
| Meas[63] | Active |
| Meas[64] | Active |
| Meas[65] | Active |
| Meas[66] | Active |
| Motion Correction | On |
| Spatial Filter | On |
| Filter Width | 4.0 mm |
| Measurements | 200 |
| Delay in TR | 0.00 ms |

### Sequence - Part 1

| | |
|---|---|
| Sequence Name | epfid |
| Excitation | Standard |
| RF Pulse Type | Normal |
| Gradient Mode | Fast* |
| Bandwidth | 2368 Hz/Px |
| Echo Spacing | 0.53 ms |
| Free Echo Spacing | Off |
| EPI Factor | 64 |

### Sequence - Part 2

| | |
|---|---|
| Introduction | Off |

**\\NRA\Schädel_32ch\Ischämie\CRPPstroke20191228_32ch_New_Patients6-\t1mprage_tra_HighRes_Nch_32ch ***

TA: 8:14 min Coil Selection: Auto Voxel Size: 0.8×0.8×1.0 mm³ Acc:: 2 Rel. SNR: 1.00

### Properties

| | |
|---|---|
| Start measurement without further preparation | On |
| Wait for User to Start | Off |
| Start measurements | Single Measurement |
| Prio Recon | Off |
| Auto Open Inline Display | Off |
| Auto Close Inline Display | Off |
| Load Images to MR View&GO | On |
| Auto Store Images | On |
| Load Images to Stamp Segments | Off |
| Load Images to Graphic Segments | Off |
| Graphic segment | Default |
| Inline Movie | Off |

### Routine

| | |
|---|---|
| Slab Group | 1 |
| Slabs | 1 |
| Distance Factor | 50 % |
| Position | L0.0 P1.6 H13.6 mm |
| Orientation | Transversal |
| Phase Encoding Dir. | R >> L |
| Slices per Slab | 176 |
| Phase Oversampling | 10 % |
| Slice Oversampling | 27.3 % |
| FoV Read | 230 mm |
| FoV Phase | 100.0 % |
| Slice Thickness | 1.0 mm |
| TR | 2200.0 ms |
| TE | 5.17 ms |
| Averages | 1 |
| Concatenations | 1 |
| AutoAlign | Head > Brain |
| Coil Elements | HEA;HEP |

### Contrast - Common

| | |
|---|---|
| TR | 2200.0 ms |
| TE | 5.17 ms |
| Magn. Preparation | Non-sel. IR |
| TI | 900 ms |
| Flip Angle | 8 deg |
| Fat-Water Contrast | Standard |
| Dark Blood | Off |
| Reconstruction | Magnitude |

### Contrast - Dynamic

| | |
|---|---|
| Dynamic Mode | Standard |
| Measurements | 1 |
| Multiple Series | Each Measurement |
| Reordering | Linear Rot. |

### Resolution - Common

| | |
|---|---|
| FoV Read | 230 mm |
| FoV Phase | 100.0 % |
| Slice Thickness | 1.0 mm |
| Base Resolution | 288 |
| Phase Resolution | 100 % |
| Slice Resolution | 100 % |
| Interpolation | Off |

### Resolution - Acceleration

| | |
|---|---|
| Acceleration mode | GRAPPA |
| Reference Scans | Integrated |
| Acceleration Factor PE | 2 |
| Reference Lines PE | 24 |
| Acceleration Factor 3D | 1 |
| Phase Partial Fourier | Off |
| Slice Partial Fourier | Off |
| Asymmetric Echo | Allowed |
| Elliptical Scanning | Off |

### Resolution - Filter

| | |
|---|---|
| Raw Filter | Off |
| Elliptical Filter | Off |
| Distortion Correction | 3D |
| Normalize | Prescan |
| Image Filter | On |

### Geometry - Common

| | |
|---|---|
| Slab Group | 1 |
| Slabs | 1 |
| Distance Factor | 50 % |
| Position | L0.0 P1.6 H13.6 mm |
| Orientation | Transversal |
| Phase Encoding Dir. | R >> L |
| Slices per Slab | 176 |
| Phase Oversampling | 10 % |
| Slice Oversampling | 27.3 % |
| FoV Read | 230 mm |
| FoV Phase | 100.0 % |
| Slice Thickness | 1.0 mm |
| TR | 2200.0 ms |
| Multi-Slice Mode | Single Shot |
| Series | Ascending |
| Concatenations | 1 |

### Geometry - AutoAlign

| | |
|---|---|
| Slab Group | 1 |
| Position | L0.0 P1.6 H13.6 mm |
| Orientation | Transversal |
| Phase Encoding Dir. | R >> L |
| AutoAlign | Head > Brain |
| Initial Position | L0.0 P1.6 H13.6 |
| R | 0.0 mm |
| P | 1.6 mm |
| H | 13.6 mm |
| Initial Orientation | Transversal |
| Initial Rotation | 91.66 deg |

### Geometry - Navigator

### Geometry - Tim Planning Suite

| | |
|---|---|
| Set-n-Go Protocol | Off |
| Table Position | 0 mm |
| Table Position | H |
| Inline Composing | Off |

### System - Miscellaneous

| | |
|---|---|
| Coil Selection | Auto Coil Select |
| MSMA | S - C - T |

### System - Miscellaneous

| | |
|---|---|
| Sagittal | R >> L |
| Coronal | A >> P |
| Transversal | F >> H |
| Coil Combination | Adaptive Combine |
| Matrix Optimization | Off |

### System - Adjustments

| | |
|---|---|
| Adjustment Strategy | Standard |
| B0 Shim | Tune up |
| B1 Shim | TrueForm |
| Adjustment Tolerance | Auto |
| Adjust with Body Coil | Off |
| Confirm Frequency | Never |
| Assume Silicone | Off |

### System - Adjust Volume

| | |
|---|---|
| Position | Isocenter |
| Orientation | Transversal |
| Rotation | 0.00 deg |
| A >> P | 263 mm |
| R >> L | 350 mm |
| F >> H | 350 mm |
| Reset | Off |

### System - Tx/Rx

| | |
|---|---|
| Frequency 1H | 123.256229 MHz |
| ? Ref. Amplitude 1H | 0.000 V |
| Reset | Off |
| Correction Factor | 1.00 |
| Image Scaling | 1.000 |

### Physio - Signal

| | |
|---|---|
| 1st Signal/Mode | None |
| TR | 2200.0 ms |
| Concatenations | 1 |

### Physio - Cardiac

| | |
|---|---|
| Fat-Water Contrast | Standard |
| Magn. Preparation | Non-sel. IR |
| TI | 900 ms |
| Dark Blood | Off |
| FoV Read | 230 mm |
| FoV Phase | 100.0 % |
| Phase Resolution | 100 % |
| Dynamic Mode | Standard |

### Physio - PACE

| | |
|---|---|
| Resp. Control | Off |
| Concatenations | 1 |

### Inline - Subtraction

| | |
|---|---|
| Subtract | Off |
| Measurements | 1 |
| StdDev | Off |
| Save Original Images | On |

### Inline - Cardiac

| | |
|---|---|
| Magn. Preparation | Non-sel. IR |
| Save Original Images | On |
| TE | 5.17 ms |
| TR | 2200.0 ms |

### Inline - MIP

| | |
|---|---|
| MIP Sag | Off |
| MIP Cor | Off |
| MIP Tra | Off |
| MIP Time | Off |
| Radial MIP | Off |
| Save Original Images | On |
| MPR Sag | Off |
| MPR Cor | Off |
| MPR Tra | Off |

### Inline - Composing

| | |
|---|---|
| Inline Composing | Off |

### Sequence - Part 1

| | |
|---|---|
| Sequence Name | tfl_r |
| Dimension | 3D |
| Excitation | Slab-sel. |
| RF Pulse Type | Fast |
| Gradient Mode | Normal |
| Flow Compensation | On |
| Reordering | Linear Rot. |
| Bandwidth | 250 Hz/Px |
| Echo Spacing | 10.18 ms |
| Asymmetric Echo | Allowed |
| Turbo Factor | 317 |

### Sequence - Part 2

| | |
|---|---|
| Introduction | On |
| RF Spoiling | On |
| Incr. Gradient Spoiling | On |

### Sequence - Assistant

| | |
|---|---|
| SAR Assistant | Off |
